# Quantifying contribution of Lipoprotein (a) to atherogenic lipoprotein burden – a novel particle-based approach

**DOI:** 10.1101/2021.06.30.21259801

**Authors:** Michael Chilazi, Weili Zheng, Jihwan Park, Francoise A. Marvel, Shireen Khoury, Steven R. Jones, Seth S. Martin

**Author notes:** **Address for Correspondence:** Seth S. Martin, MD, MHS, FACC, FAHA, FASPC, Associate Professor of Medicine (Cardiology), Epidemiology, and Health Informatics Director, Advanced Lipid Disorders Program, Ciccarone Center for the Prevention of Cardiovascular Disease, 600 N. Wolfe Street, Carnegie 591, Baltimore, MD, 21287, Facsimile, Twitter: @SethShayMartin. **Disclosures:** SSM has research support from the American Heart Association (20SFRN35380046 and COVID19-811000), PCORI (ME-2019C1-15328), National Institutes of Health (P01 HL108800), the Aetna Foundation, the David and June Trone Family Foundation, the Pollin Digital Innovation Fund, the PJ Schafer Cardiovascular Research Fund, Sandra and Larry Small, CASCADE FH, Apple, Google, and Amgen. He has served as a consultant to Akcea, Amgen, AstraZeneca, Esperion, Kaneka, Novo Nordisk, Quest Diagnostics, Regeneron, REGENXBIO, Sanofi, and 89bio. He is a co-inventor on a system to estimate LDL cholesterol levels, patent application pending.

## Abstract

**Background:** Elevated lipoprotein (a) [Lp(a)] is an independent risk factor for atherosclerotic cardiovascular disease (ASCVD). As clinical LDL cholesterol [LDL-C] incorporates cholesterol from Lp(a) [Lp(a)-C], there is interest in quantifying the contribution of Lp(a)-C to LDL-C given implications for risk assessment, diagnosis, and treatment. Estimating Lp(a)-C is subject to inaccuracies; measuring Lp(a) particle number [Lp(a)-P] is more accurate.

**Objective:** To capture how Lp(a) contributes to the atherogenic lipoprotein burden, we demonstrate a particle-based approach using readily available measures of Lp(a)-P and apolipoprotein B (apoB).

**Methods:** Using the Very Large Database of Lipids (VLDbL), we compared Lp(a)-P (nmol/L) with all atherogenic particles (“non-HDL-P”). Non-HDL-P was calculated by converting apoB mass to molar concentration using the preserved molecular weight of apoB100 (512 kg/mol). We calculated the percentage of Lp(a)-P relative to non-HDL-P by Lp(a)-P deciles and stratified across sex, age, triglycerides, LDL-C, and non-HDL-C.

**Results:** 158,260 patients from the VLDbL were included. The fraction Lp(a)-P/non-HDL-P increased with rising Lp(a)-P. Lp(a)-P comprised on average 3% of atherogenic particles among the study population and 15% at the highest Lp(a)-P decile. Findings were similar when stratified by sex. When stratified by age, Lp(a)-P/non-HDL-P was highest among the youngest and oldest patients. Lp(a)-P/non-HDL-P decreased at higher levels of triglycerides and LDL-C owing to larger contributions from VLDL and LDL.

**Conclusions:** We demonstrate a particle-based approach to quantify the contribution of Lp(a) to total atherogenic particle burden using validated and widely available laboratory assays. Future research applying this method could define clinically meaningful thresholds and inform use in risk assessment and management.

## Introduction

Evidence from cohort and genetic studies have solidified the role of Lp(a) in mediating ASCVD, now recognized in the American Heart Association/American College of Cardiology lipid guidelines as a risk-enhancing factor.^1^ LDL-C on conventional lipid panels includes the cholesterol component of Lp(a), as LDL-C is actually the sum of the cholesterol within LDL, IDL, and Lp(a). There has been considerable interest in quantifying the contribution of Lp(a)-C to LDL-C due to implications for risk assessment, diagnosis, and treatment. With novel Lp(a)-lowering therapies in development,^2^ it will become increasingly valuable to understand how Lp(a) changes relative to LDL and other non-HDL lipoproteins.

There is presently no commercially available assay to accurately measure Lp(a)-C, although promising monoclonal antibody strategies are in development.^3^ We previously showed that commonly used conversion factors to estimate Lp(a)-C from total mass or particle number led to inaccuracy at clinically meaningful Lp(a)-C levels.^4^ Fortunately, standardized approaches to measure Lp(a) particle number have been developed which are less prone to inaccuracies of mass measurement.^5^ To address the need to quantify Lp(a)’s contribution to the lipoprotein burden, we propose a particle-based approach illustrating how Lp(a) contributes not just to LDL-C, but more broadly to all atherogenic particles (i.e., non-HDL apoB-containing particles, herein referred to as non-HDL-P).

This approach is possible because each atherogenic particle carries one apoB100 molecule, the molecular weight of which is conserved across individuals, allowing for accurate calculation of particle number by converting apoB mass (mg/dL) to molar concentration (nmol/L).^6,7^ While apoB includes both apoB100 and apoB48 (found on intestinal chylomicrons), the concentration of apoB48 is usually negligible.^8^ Here, we perform a descriptive analysis using the Very Large Database of Lipids (VLDbL) to illustrate the feasibility of this particle-based method to compare Lp(a)-P with all atherogenic particles using commercially available measurements.

## Methods

### Study Population

Data were extracted from the Very Large Database of Lipids (VLDbL). The VLDLbL is registered at http://www.clinicaltrials.gov (NCT01698489). The data were obtained from Vertical Auto Profile (VAP) Diagnostics Laboratory (Birmingham, AL) from 2006-2015 and transferred to the academic investigators under a data use agreement.^9^ The majority of specimens were sent from primary care clinics (∼85%) with the remainder originating from inpatient settings and specialty centers. Lipid distributions within the VLDbL mirror those observed in the National Health and Nutrition Examination Survey, a representative population-based survey.^9^ We analyzed all samples from the second harvest of the VLDbL and all individuals with Lp(a)-P (nmol/L) and apoB (mg/dL) values were included in the study. The Johns Hopkins Institutional Review Board declared our study exempt, which used de-identified data routinely collected from a commercial laboratory.

### Lipid Measurements

The VLDbL contains direct measurements of the cholesterol components of all major lipoproteins including LDL, IDL, VLDL, HDL, and Lp(a). VLDbL employs Vertical Auto Profile ultracentrifugation to separate lipoproteins into various classes and the cholesterol component is subsequently measured via spectrophotometric absorbance.^10^ The accuracy of VAP measurements has been previously validated against other reference laboratories.^9,11^ In addition to cholesterol measurements, VLDbL also contains secondary analytes, including apoB, Lp(a) particle number, and high-sensitivity CRP, amongst others. Lp(a)-P in the VLDbL was measured via the Denka-Seiken immunoassay which correlates with isoform-insensitive ELISA tests owing to its use of a five-point calibration system traceable to the WHO/IFCC Lp(a) reference material.^5^ ApoB in VLDbL was measured by a World Health Organization standardized immunoassay.^12^

### Statistical Analysis

The mass of apoB was converted to molar concentration using the preserved and previously established molecular weight of apoB100 of approximately 512 kg/mol.^6,7^ Of note, there exists in the literature a range of apoB100 molecular weights spanning 512-515 kg/mol depending on the employed methodlogy.^13^ Given this narrow range, the effects on apoB concentration is minimal, and for the purposes of this study, 512 kg/mol was chosen as it was cited in the foundational studies. Molar concentration of atherogenic particles (non-HDL-P) was expressed in nmol/L to enable direct comparison with Lp(a)-P. The study population was divided into deciles with increasing Lp(a)-P concentrations. Mean and median percentage of Lp(a)-P/non-HDL-P were subsequently calculated for each decile. Results were further stratified by sex, age, triglyceride, LDL-C, and non-HDL-C levels. Statistical analysis was performed with Stata, version 16.0.

## Results

Overall, 158,260 participants from the VLDbL met inclusion criteria. Median age was 57 years (IQR: 45-68 years). 58% of the study population was female. Complete demographic information is illustrated in Table 1.

**Table 1.** Patient Demographics

|  | Study population<br>(n=158,260) |
| --- | --- |
| Age, yr, median (IQR) | 57 (45-68) |
| Sex, n (%) |  |
| Male | 65961 (42%) |
| Female | 91334 (58%) |
| Lipid values, mg/dL, median (IQR) |  |
| Total Cholesterol | 198 (169-230) |
| HDL-C | 52 (43-64) |
| Triglycerides | 115 (82-168) |
| Measured LDL-C | 117 (93-144) |
| LDL-C<sub>Friedewald</sub> | 115 (91-143) |
| LDL-C<sub>Martin/Hopkins</sub> | 117 (93-144) |
| Measured Lp(a)-C | 7 (5-11) |
| Estimated Lp(a)-C | 6 (3-16) |
| Lp(a)-P, nmol/L | 47 (21-124) |
| ApoB (mg/dL) | 92 (77-109) |
| non-HDL-P, nmol/L | 1797 (1504-2129) |
| non-HDL-P minus Lp(a)-P, nmol/L | 1708 (1412-2041) |
| Lipid ratios, median (IQR) |  |
| Lp(a)-P : non-HDL-P | 0.027 (0.012-0.068) |
| Estimated Lp(a)-C : LDL-C<sub>Friedewald</sub> | 0.054 (0.023-0.134) |

Mean percentage of Lp(a)-P relative to non-HDL-P is depicted in Figure 1. The relative contribution of Lp(a)-P to the total atherogenic population varied significantly across the study group according to baseline Lp(a)-P concentration. Individuals in the lowest decile, with Lp(a)-P ranging from 0.2 to 12.4 nmol/L, had a mean percentage of 0.62% (median 0.59%; IQR 0.49-0.73%), illustrating a negligible makeup of their total atherogenic particles by Lp(a)-P. This is in contrast to individuals in the highest decile with Lp(a)-P concentrations of 195 nmol/L or higher where the mean percentage was nearly 15% (median 13.7%; IQR 11.0-17.5%), representing a more sizable contribution of Lp(a)-P to the total atherogenic lipoprotein concentration. Median percentage of Lp(a)-P/non-HDL-P for the whole study population was 3% (IQR 1-7%). Nearly 40% of participants saw their Lp(a)-P make up 5% or more of their total atherogenic burden.

**Figure 1.**
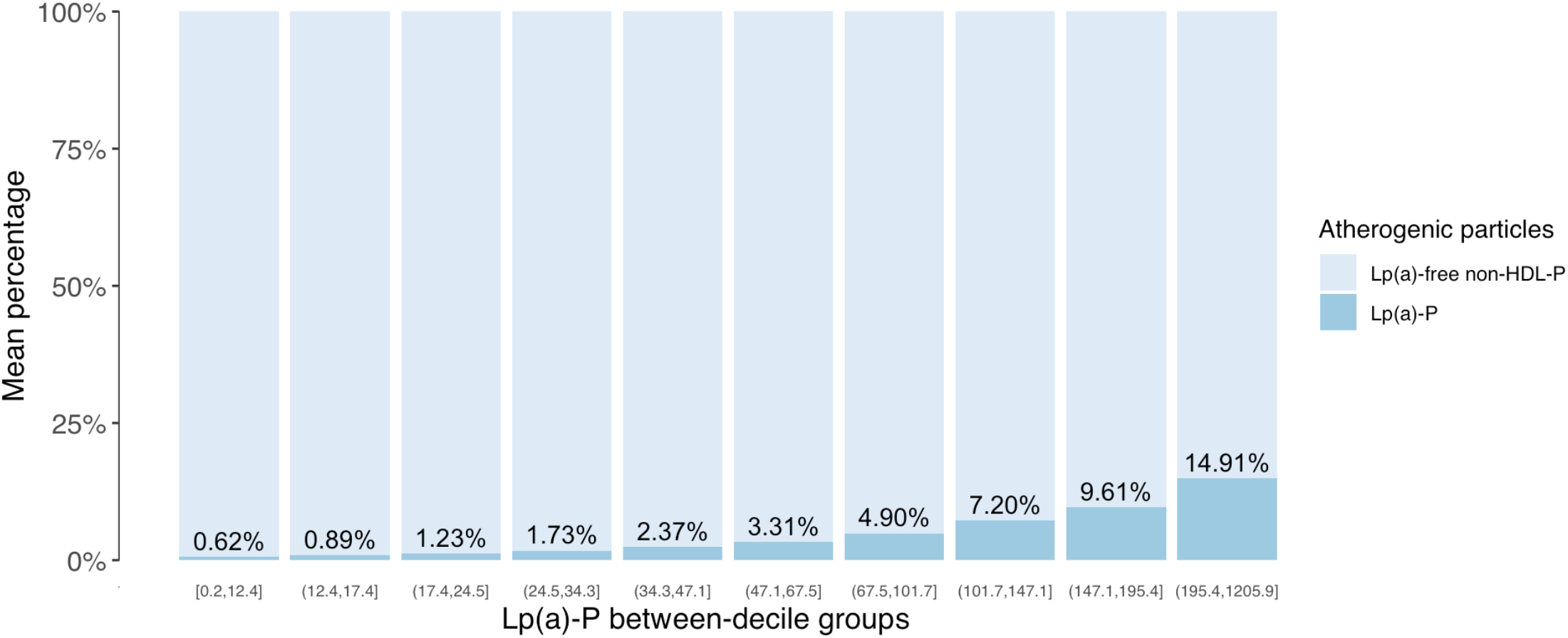
Mean percentage of Lp(a)-P relative to the total number of atherogenic particles (non-HDL-P), stratified into deciles with increasing Lp(a)-P concentration. Range of Lp(a)-P within each decile noted on x-axis.

Among individuals with higher Lp(a)-P concentrations, Lp(a) made greater contributions to their atherogenic population. That the percentage of Lp(a)-P/non-HDL-P did not remain constant suggests that high Lp(a)-P among these individuals was not accompanied by proportional increases in other atherogenic particles contributing to non-HDL-P (e.g., LDL, IDL, VLDL).

Percentage of Lp(a)-P/non-HDL-P was further stratified by sex, age, triglycerides, LDL-C, and non-HDL-C. When stratified by sex, percentages across Lp(a)-P deciles were equivalent between men and women (Figure 2). Median percentage by Lp(a)-P decile paralleled those seen in the overall population.

**Figure 2.**
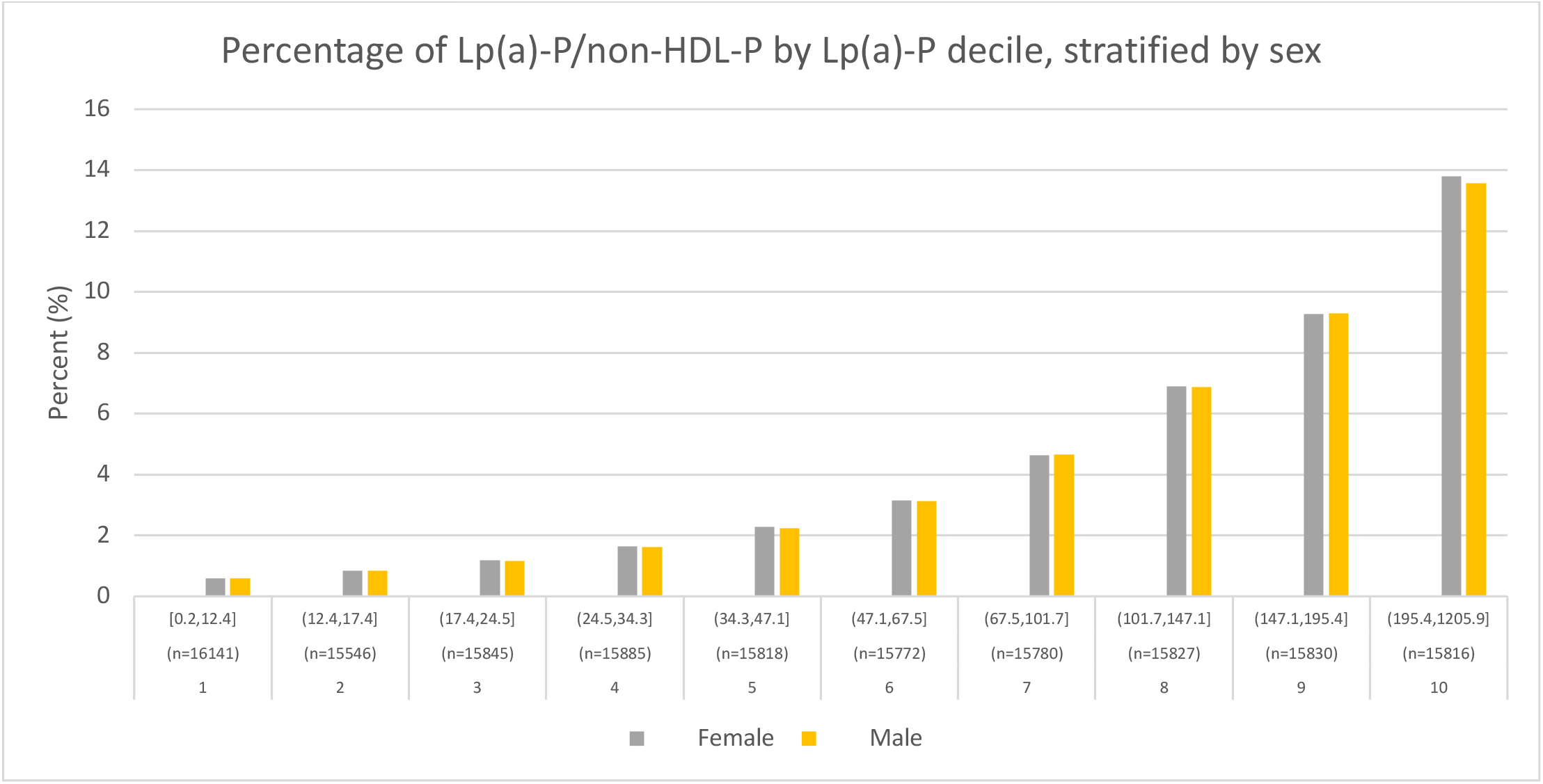
Median Lp(a)-P/non-HDL-P organized by increasing Lp(a)-P decile (range of Lp(a)-P and sample size within each decile noted), stratified by sex.

When stratified by age, percentage of Lp(a)-P/non-HDL-P demonstrated a bimodal distribution. Within each Lp(a)-P decile, higher percentages were observed at the extreme ends of the age spectrum (< 20 years old and > 70 years old, respectively). The lowest percentages were seen consistently in the age range of 40-49. (Supplementary Figure 1) To determine the significance of this degree of difference between age groups, we calculated marginal mean percentages for each group, which allows for adjustment of the covariate, here being Lp(a)-P decile. Using a beta regression and a pairwise comparison, we found that the mean percentages were significantly different across all age groups except between groups aged 20-29 and 70 or older (Supplementary Figure 2).

When stratified by triglyceride levels, higher triglycerides were associated with reductions in the percentage of Lp(a)-P/non-HDL-P across Lp(a)-P deciles. As noted previously, non-HDL incorporates VLDL, the primary carrier of circulating triglycerides. On average, the percentage of Lp(a)-P/non-HDL-P dropped by 25% when triglycerides crossed a threshold of 400 or greater (Figures 3 and 4).

**Figure 3.**
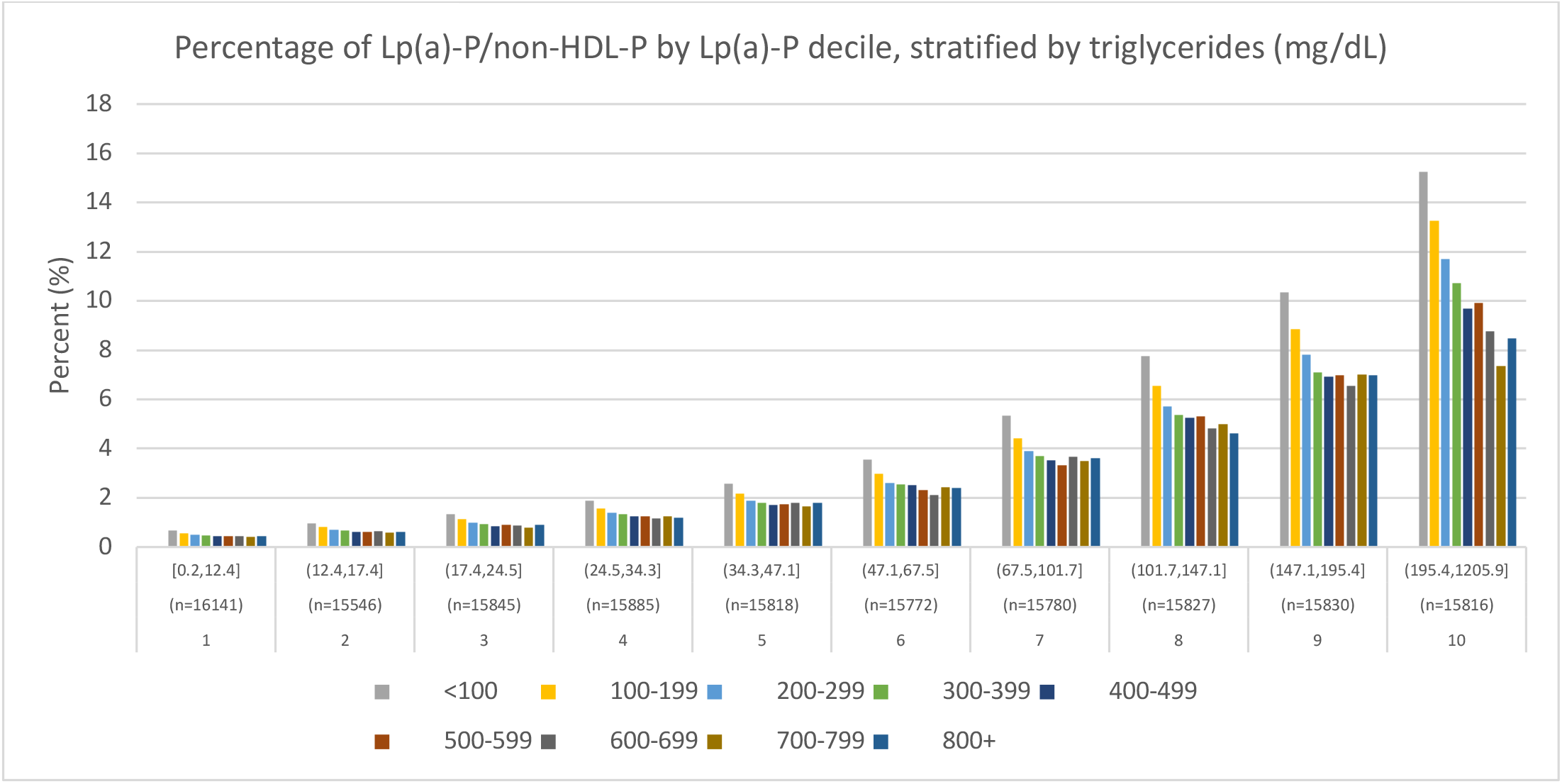
Median Lp(a)-P/non-HDL-P organized by increasing Lp(a)-P decile (range of Lp(a)-P and sample size within each decile noted), stratified by TG levels.

**Figure 4.**
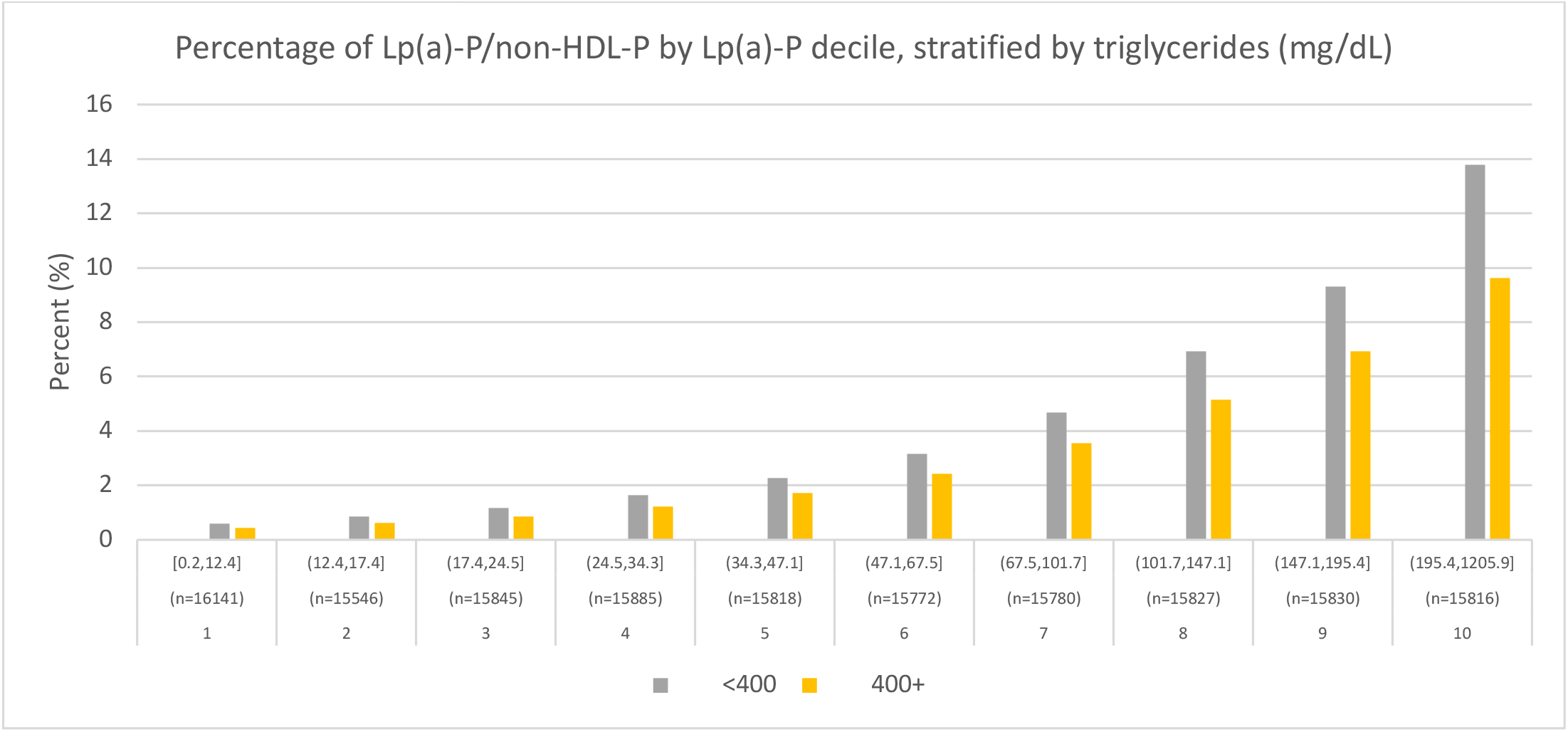
Median Lp(a)-P/non-HDL-P organized by increasing Lp(a)-P decile (range of Lp(a)-P and sample size within each decile noted), stratified by TG levels less than or greater than or equal to 400.

When stratified by LDL-C, we found a similar relationship seen with triglycerides whereby across all Lp(a)-P deciles, the percentage of Lp(a)-P relative to non-HDL-P decreased with increasing LDL-C, likely explained by LDL making a more significant footprint on the atherogenic lipoprotein burden. (Figure 5) There was generally a reduction of ≥ 50% in Lp(a)-P/non-HDL-P when comparing individuals with LDL-C < 50 versus those > 200 mg/dL. As expected, the percentage of Lp(a)-P/non-HDL-P also declined with rising non-HDL-C levels (Figure 6), keeping in line with the relationship seen with rising LDL and VLDL which both contribute to non-HDL-C.

**Figure 5.**
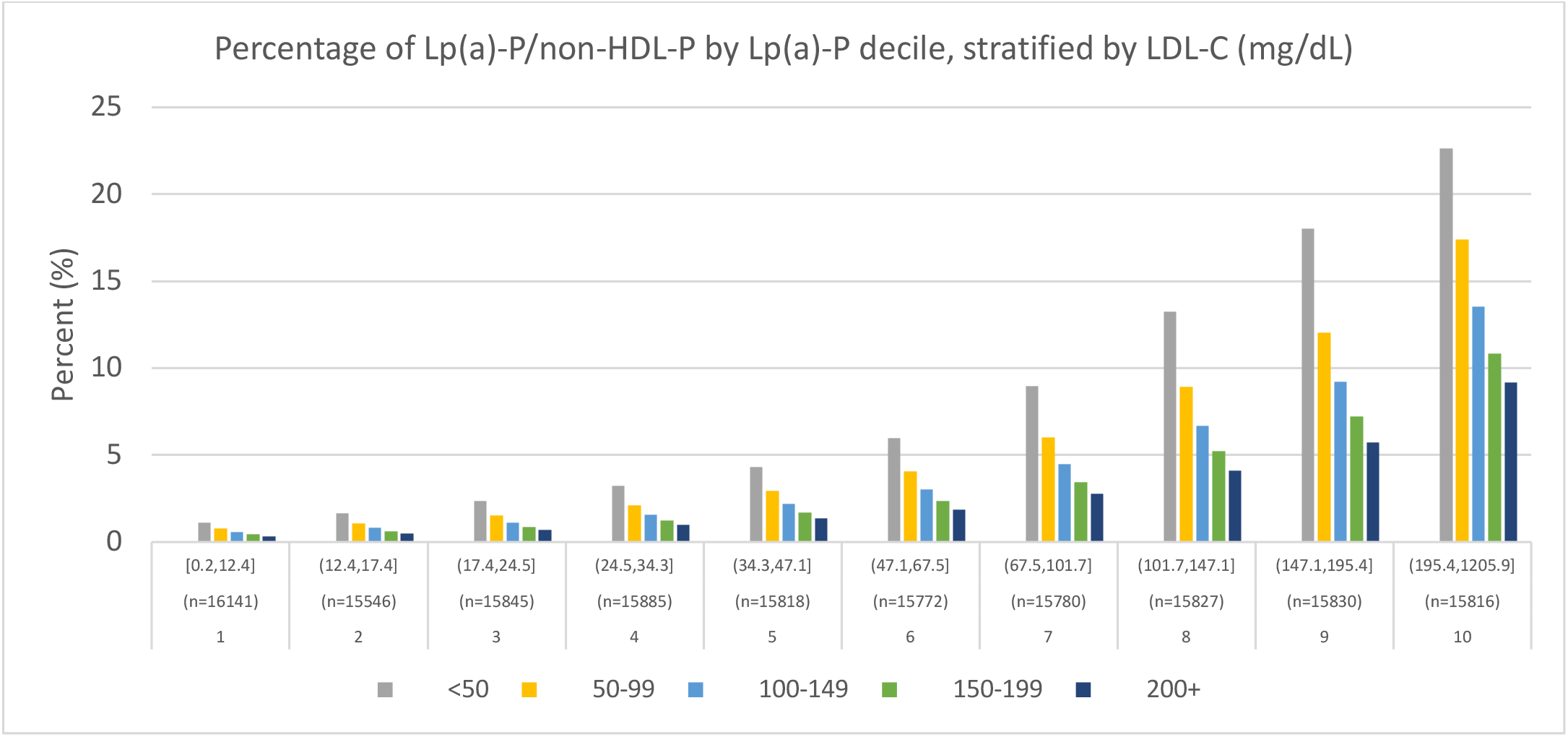
Median Lp(a)-P/non-HDL-P organized by increasing Lp(a)-P decile (range of Lp(a)-P and sample size within each decile noted), stratified by LDL-C.

**Figure 6.**
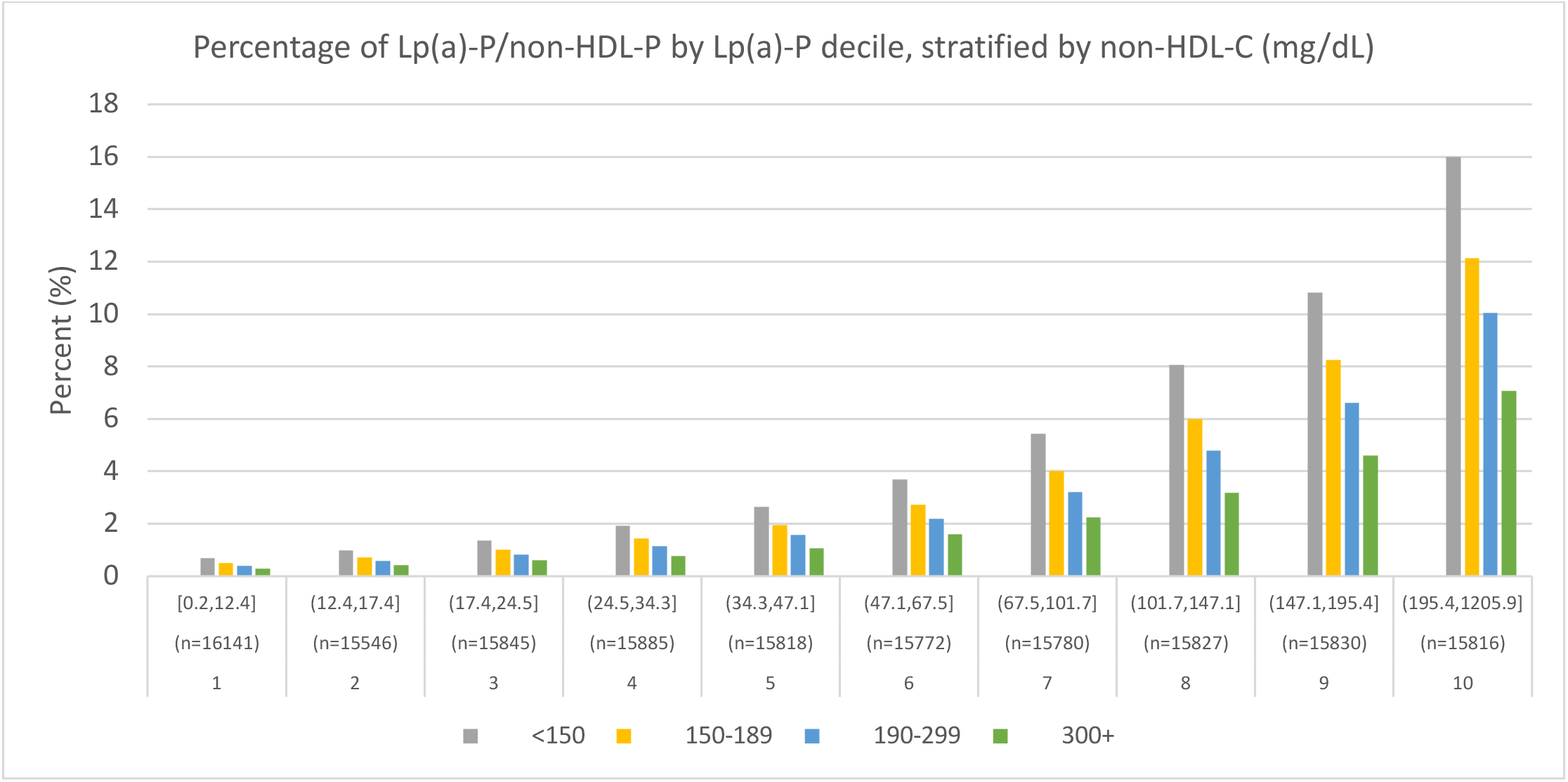
Median Lp(a)-P/non-HDL-P organized by increasing Lp(a)-P decile (range of Lp(a)-P and sample size within each decile noted), stratified by non-HDL-C.

## Discussion

This descriptive analysis demonstrates the simplicity and feasibility of quantifying the relative contribution of Lp(a)-P to the overall concentration of atherogenic particles, employing two commercial, widely available and clinically used assays for apoB and Lp(a)-P. The use of this particle-based approach provides an alternative method circumventing inaccuracies associated with mass estimation and expands understanding of how Lp(a) compares not just with LDL but with all atherogenic particles.^14^

We demonstrated that individuals with higher baseline Lp(a)-P concentrations had greater contribution from Lp(a) to their total atherogenic particles. When stratifying by triglyceride and LDL-C levels, the percentage of Lp(a)-P/non-HDL-P decreased with rising triglycerides and LDL-C, likely driven by the greater contribution of VLDL and LDL respectively to the atherogenic lipoprotein burden. When stratifying by age, we observed that percentages of Lp(a)-P/non-HDL-P were highest at the extremes of age. We confirmed statistical significance between the youngest and oldest patients compared to the remaining cohort. This observation may be explained by more favorable lipid profiles seen among the youngest and oldest participants. Particularly for patients surviving to age 70 and beyond, survivor bias may contribute to more favorable lipid profiles. Because Lp(a) is genetically determined, it likely makes a more prominent atherogenic contribution among patients with otherwise well-controlled LDL-C and triglycerides.^15^ Moreover, the use of LDL and triglyceride lowering therapies may be higher among older age groups, leading to greater contributions from Lp(a) to their residual atherogenic burden.

Individuals with greater Lp(a)-P had higher Lp(a)-P/non-HDL-P ratios, which suggests that their Lp(a)-P levels were not accompanied by proportional increases in other atherogenic particles. This too is likely explained by the strong genetic determination of Lp(a) concentration.^16^ Lp(a) levels are less affected by environmental factors such as diet and exercise as compared to LDL and VLDL.^17,18^ The strong genetic influence on Lp(a) and the meaningful contribution Lp(a)-P can make to the atherogenic burden as seen in our study --up to 15% in the highest decile --reinforce the need for targeted Lp(a) therapies for high-risk patients.

The need for Lp(a) therapies is especially salient given that the majority of dyslipidemia therapies operate via LDL and triglyceride reductions with minimal effect on Lp(a). As a result, treatment effects would be expected to result in an increasing percentage of Lp(a)-P/non-HDL-P. An important area for future investigation is how, following conventional lipid lowering treatments, increases in this Lp(a) fraction might predict residual ASCVD risk. An elevated Lp(a) fraction may identify Lp(a) as an appropriate therapeutic target after adequate control of LDL and triglycerides to mitigate residual risk. Importantly, clinical interpretration of the Lp(a)-P/non-HDL-P and meaningful thresholds may vary between treated and untreated individuals. In contrast to conventional lipid lowering therapies, antisense oligonucleotide therapies currently in development, which directly reduce the production of Lp(a), would be expected to lead to significant reductions in Lp(a)-P/non-HDL-P.^2^

An important consideration when comparing Lp(a)-P to non-HDL-P is that the risk conferred by a single Lp(a) particle likely differs from that of LDL or VLDL. While there is evidence to suggest that LDL and VLDL particles carry equivalent atherogenic risk,^19^ the same may not be true for Lp(a). Lp(a) contains all the atherogenic potential of LDL due its similar cholesterol-core, but the associated apo(a) moiety may confer additional risk secondary to pro-inflammatory and pro-atherogenic properties. Owing to its oxidized phospholipids and lysine residues mediating adherence to the intimal wall, apo(a) accelerates the inflammation driving plaque formation. Moreover, homology of apo(a) with plasminogen is suggested to interfere with thrombolysis, conferring additional prothrombotic potential.^14^

These unique properties of Lp(a) likely contribute to residual atherosclerotic risk seen even after LDL-C is aggressively reduced to levels below 70 mg/dL.^20,21^ High-risk patients are defined by AHA/ACC guidelines as those with Lp(a) concentrations greater than 125 nmol/L.^1^ In our study, at these particle concentrations, Lp(a) made up 7% or more of the total atherogenic burden. That Lp(a) can mediate risk when only representing 7% of total atherogenic particles is a testament to its atherosclerotic potency. These additional mechanisms through which Lp(a) drives atherosclerosis should be considered when interpreting percentages of Lp(a)-P/non-HDL-P; Lp(a)’s contribution towards total risk may be higher than the raw percentage suggests.

Further research will be required to define clinically meaningful thresholds for the percentage of Lp(a)-P/non-HDL-P. Current AHA/ACC guidelines employ Lp(a)-P levels of 125 nmol/L or greater to identify high-risk patients.^1^ This level of Lp(a)-P was crossed in the 8^th^ decile of our study population where the mean and median percentage of Lp(a)-P/non-HDL-P was 7.2% and 6.9%, respectively. This finding may identify > 7% as a clinically meaningful threshold of Lp(a)-P relative to the total atherogenic population, of which nearly 30% of our study population crossed, however further investigation is needed. One would anticipate that clinical significance of Lp(a) depends on the absolute concentration of Lp(a) in addition to its percentage contribution to non-HDL-P.

## Limitations

Because the VLDbL does not contain data regarding clinical outcomes, the descriptive analysis performed here cannot assess the performance of Lp(a)-P/non-HDL-P in predicting risk of adverse cardiovascular events. VLDbL also does not contain treatment information and therefore changes in Lp(a)-P/non-HDL-P before and after therapy could not be investigated. Society guidelines for dyslipidemia largely employ cholesterol mass to define treatment targets. Until further population studies are performed applying particle-based metrics such as Lp(a)-P/non-HDL-P, we recognize that this percentage will only provide clinicians with a qualitative sense of the residual risk posed by Lp(a) in the assessment and management of dyslipidemia.

## Conclusion

We presented a novel method to quantify Lp(a)’s contribution to the total atherogenic lipoprotein burden by employing two measurements used in clinical practice and with readily available commercial assays --apoB (mg/dL) and Lp(a)-P (nmol/L). This method fulfills a need to deconstruct total atherosclerotic risk into its component parts to better inform risk assessment and treatment targets. A particle-based approach maintains accuracy by avoiding reliance on mass estimations. Further investigation is needed to define clinically meaningful thresholds for Lp(a)-P/non-HDL-P that will inform risk stratification and management.

## Data Availability

The datasets used and/or analyzed during the current study are available from the corresponding author on reasonable request.

## Supplemental Appendix

**Supplementary Figure 1.**
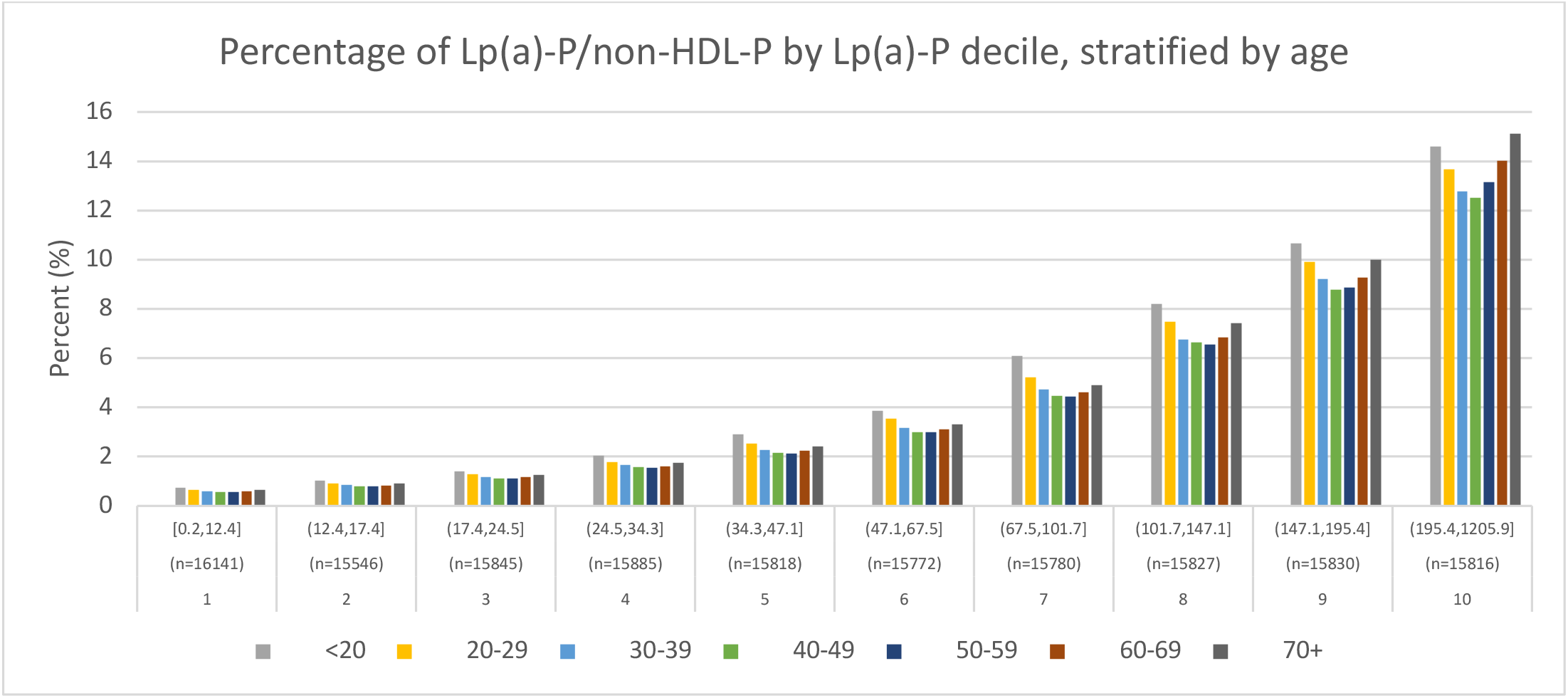
Median Lp(a)-P/non-HDL-P organized by increasing Lp(a)-P decile (range of Lp(a)-P and sample size within each decile noted), stratified by age.

**Supplementary Figure 2.**
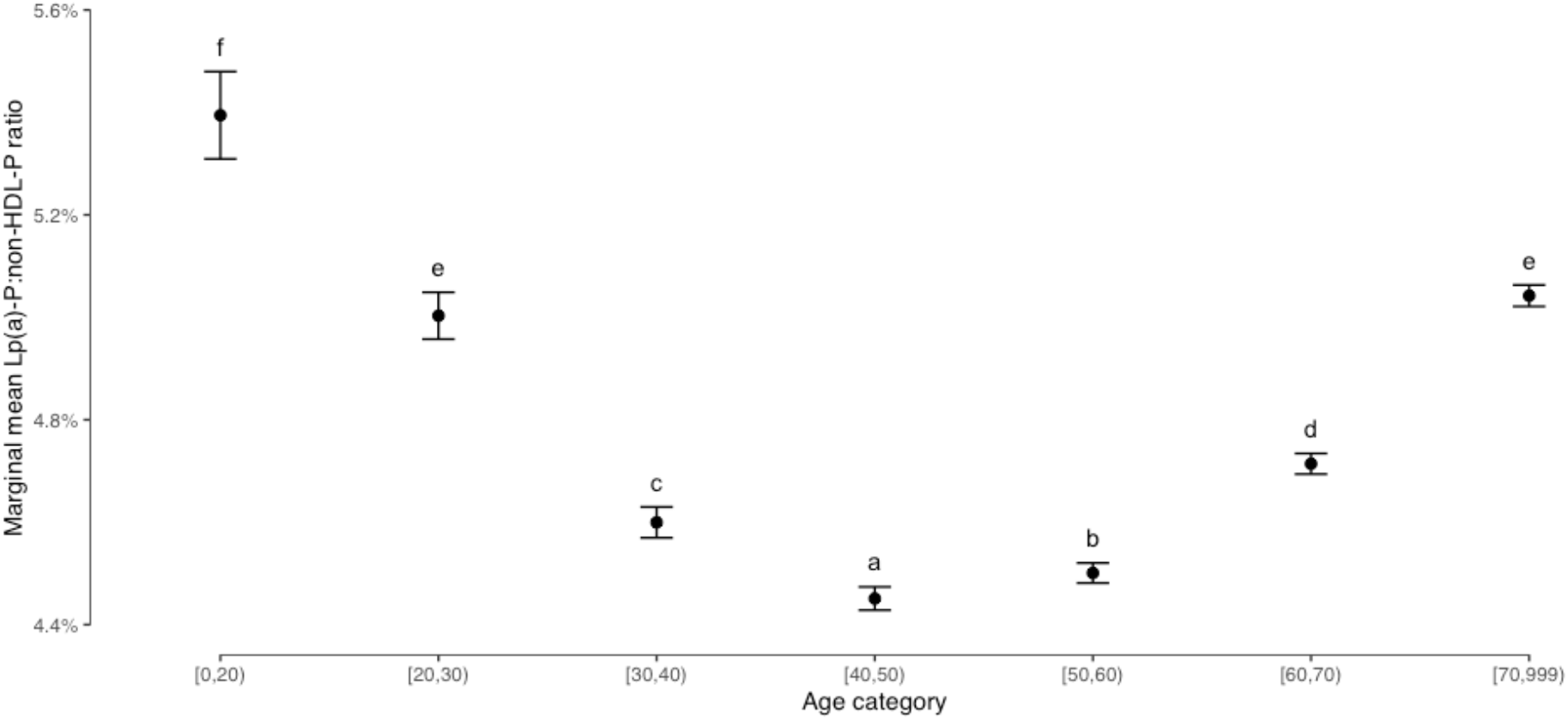
Marginal mean of Lp(a)-P/non-HDL-P by age category. Marginal means with unique letters denote statistical significance relative to other mean values.

